# Unscreenable: The Burden, Structure, and Analytic Consequences of "Unable to Assess" Delirium Documentation in the Intensive Care Unit

**DOI:** 10.64898/2026.06.13.26355598

**Authors:** Alon Gorenshtein, Yosef Adiniaev, Mahmud Omar, Yiftach Barash, Eyal Klang, Oved Daniel

## Abstract

**Objective:** To quantify the burden, structure, and downstream analytic consequences of "Unable to Assess" (UTA) delirium documentation in the intensive care unit (ICU).

**Design:** Retrospective cross-sectional and repeated-measures study.

**Setting:** A single US academic medical center (Medical Information Mart for Intensive Care IV [MIMIC-IV], 2008-2019).

**Patients:** 72,944 adult ICU stays with at least 1 delirium screen.

**Interventions:** None.

**Measurements and Main Results:** Among 610,632 screens, 130,455 (21.4%; 95% CI, 21.0%-21.8%) were recorded as UTA, exceeding the 119,052 (19.5%) scored positive. The UTA fraction rose from 2.0% at a Richmond Agitation-Sedation Scale (RASS) score of 0 to 97.8% at RASS -4; 22.0% of UTA screens occurred in arousable patients, where UTA was associated with mechanical ventilation (odds ratio [OR], 3.43; 95% CI, 3.17-3.71) and non-English primary language (OR, 3.74; 95% CI, 3.43-4.08). Building the delirium label three ways from the same patients shifted prevalence modestly (32.1% to 30.8%) and prediction (area under the curve, 0.737 to 0.719) but most affected the delirium-mortality association: in a baseline-adjusted model the OR was 4.12 (95% CI, 3.88-4.36) under complete-case handling and fell to 2.16 (95% CI, 2.06-2.27) when UTA was recoded as negative. UTA was recoverable from the observed clinical state (area under the curve, 0.95).

**Conclusions:** In this ICU cohort, Unable to Assess was the most common recorded delirium result other than Negative, exceeding positive screens; recoding it as negative roughly halved the apparent delirium-mortality association by relabeling deeply sedated, high-mortality patients. Delirium datasets should preserve and report UTA, whose concentration among arousable non-English-speaking patients is a measurable equity target.

## Introduction

Delirium is among the most common acute brain dysfunctions in critically ill adults and is associated with longer ventilation, longer hospitalization, higher mortality, and worse long-term cognition.^1,2^ Guidelines therefore recommend screening every intensive care unit (ICU) patient at least once per nursing shift with a validated instrument, most often the Confusion Assessment Method for the ICU (CAM-ICU).^3,4^ The resulting data anchor delirium prevalence estimates, prognostic studies, quality benchmarks, and a growing number of machine-learning prediction models, all of which assume that a charted screen reports the patient’s delirium status.^5^

That assumption holds only when the patient can actually be examined. The CAM-ICU and similar tools require a minimum level of arousal and the ability to follow simple commands;^4,6^ in the comatose or deeply sedated patient the instrument is not applicable, and the nurse records "Unable to Assess" rather than positive or negative. This third state is neither a delirium finding nor a conventional missing value but a documented statement that the bedside brain examination could not be performed. How often it occurs, in whom, and what happens to downstream estimates when it is dropped or recoded have not been quantified at scale. Prior work is limited to single-center audits of whether individual entries were clinically appropriate,^7^ and prediction studies routinely discard these records as a complete-case convenience.^5^

We treated the unassessable screen not as missing data to clean away but as a measurement state whose handling changes what secondary analyses conclude. Using 610,632 delirium screens from 72,944 ICU stays in a large public critical-care database,^8,9^ we quantified the burden of "Unable to Assess" documentation, separated screens unassessable by protocol from those left unassessed in arousable patients, mapped its concentration across acute brain-injury phenotypes, ICU types, and patient language, and measured the consequence of three common handling strategies for delirium prevalence, the delirium-mortality association, and prediction-model discrimination. We then asked how recoverable unassessability is from the observed clinical state.

## Methods

### Study Design and Setting

This retrospective, cross-sectional and repeated-measures study used routinely collected delirium-screening documentation from the Medical Information Mart for Intensive Care IV (MIMIC-IV) version 3.1,^8,9^ a de-identified database from the ICUs of a single US academic medical center, 2008-2019. Reporting follows the STROBE and RECORD guidelines,^10,11^ and the prediction probe follows TRIPOD-AI.^12^ Effect estimates are reported as associations; no causal claim is made.

### Cohort Identification

All adult ICU stays (≥18 years; 94,458) were eligible; the analytic cohort comprised the 72,944 with at least one charted delirium screen, and unscreened stays were retained as a denominator for the never-screened proportion.

### Data Sources and Variables

Delirium screens were identified by the "Delirium assessment" charted item (itemid 228332; Negative, Positive, or Unable to Assess [UTA]). Arousal was indexed with the Richmond Agitation-Sedation Scale (RASS, itemid 228096; -5, unarousable, to +4, combative),^6^ matched to the nearest reading within 2 hours; the matched RASS preceded the screen for 99.6% of screens. Sedation, neuromuscular blockade, ventilation, acute brain-injury phenotype, ICU type, demographics, payer, and primary language were extracted from structured tables (eMethods, eTables 1-2). The CAM-ICU feature items, which separately record an "Unable to Assess" response, were used for construct validity.

### Delirium Screen Classification

Each screen was labeled Negative, Positive, or UTA. UTA screens were split by matched RASS into structurally unassessable (RASS -4 or -5, where the CAM-ICU does not apply) and arousable (RASS -3 or higher). Because RASS -3 is borderline, potentially avoidable non-assessment was defined conservatively as UTA at RASS -2 or higher.

### Outcomes

The primary measure was the proportion of screens recorded as UTA, at the screen, patient-day, and stay level. Secondary measures were its composition by arousal and its distribution across phenotype, ICU type, and language. Handling effects were tested on delirium prevalence, the delirium-mortality association, and prediction-model discrimination. Available outcomes were in-hospital mortality, ICU length of stay, and disposition; MIMIC-IV has no structured neurologic severity scale.

### Statistical Analysis

Proportions carry Wilson 95% confidence intervals (CIs); screen-level proportions also carry patient-clustered bootstrap CIs. Adjusted associations used cluster-robust logistic regression by patient, reporting odds ratios (ORs) with 95% CIs. UTA among arousable screens was modeled with patient and process covariates; language and payer were added to race and ethnicity models to test attenuation and independent equity effects. Three handling strategies, complete-case, UTA-as-Negative, and missingness-aware, were compared for prevalence, the delirium-mortality OR, and out-of-fold area under the receiver operating characteristic curve. The primary delirium-mortality model adjusted for age, sex, the Charlson Comorbidity Index,^22^ and any acute brain injury; a prespecified secondary model added mechanical ventilation, vasopressor use, and log length of stay, which lie on the delirium-to-death path and are reported only as a sensitivity. P values used Benjamini-Hochberg correction; sensitivity analyses are in the eMethods. Analyses used Python 3.9.

### Unscreenability Probe

The probe predicted the screen-level UTA label from the observed clinical state (RASS, sedation, neuromuscular blockade, ventilation, hour, ICU type, and phenotype) by patient-grouped 5-fold cross-validation (AUROC and Brier score; eMethods). Because the features are close to definitional, it is a measurement instrument for the missingness mechanism, not a clinical prediction tool, and was not externally validated.

### Ethics

MIMIC-IV is a de-identified database released under the Health Insurance Portability and Accountability Act Safe Harbor standard; its collection and research use were approved by the institutional review boards of the Beth Israel Deaconess Medical Center and the Massachusetts Institute of Technology, with a waiver of informed consent. This secondary analysis was performed under credentialed PhysioNet access and required no additional review. Analysis code, including the eICU external-validation script (60_eicu_mechanism.py), will be made publicly available at https://github.com/Alon-Gorenshtein/study_delirium_unscreenable (archived on Zenodo on publication); eICU-CRD v2.0 is available through PhysioNet.

## Results

### Cohort

Of 94,458 adult ICU stays, 72,944 (77.2%) from 51,987 patients had at least one delirium screen and formed the analytic cohort (610,632 screens; median 5 per stay [IQR, 3-9]); the remaining 21,514 (22.8%) were never screened. Median age was 65 years, 44.0% were women, 10.0% had a primary language other than English, and 19.2% an acute brain injury; in-hospital mortality was 12.1% (Table 1). Screening rose over the period (43% of stays in 2008-2010 to 97% in 2017-2019), so the never-screened group partly reflects staged CAM-ICU adoption; even among stays of at least 24 hours, 20.6% were never screened (eTable 10).

**Table 1.**
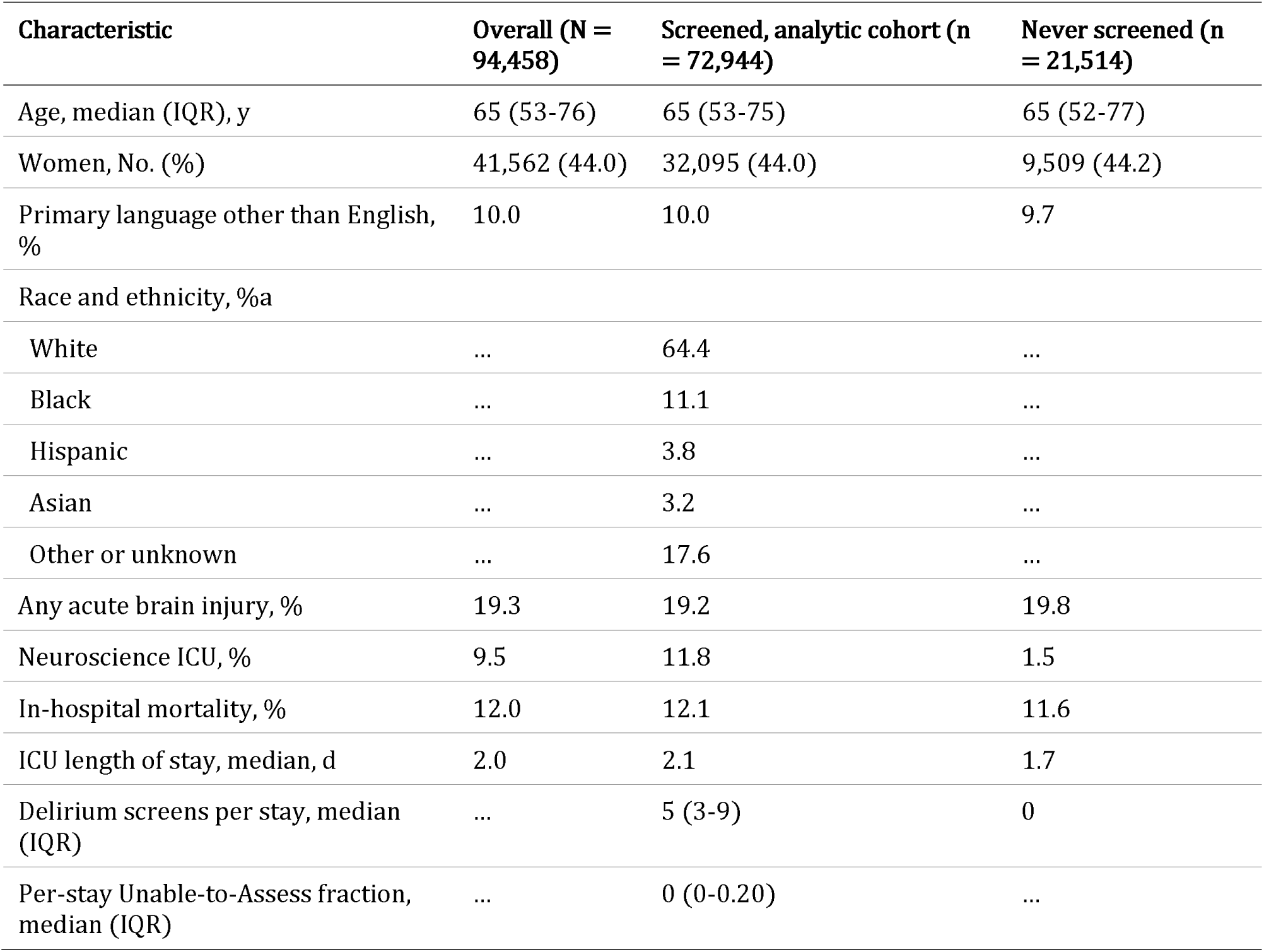
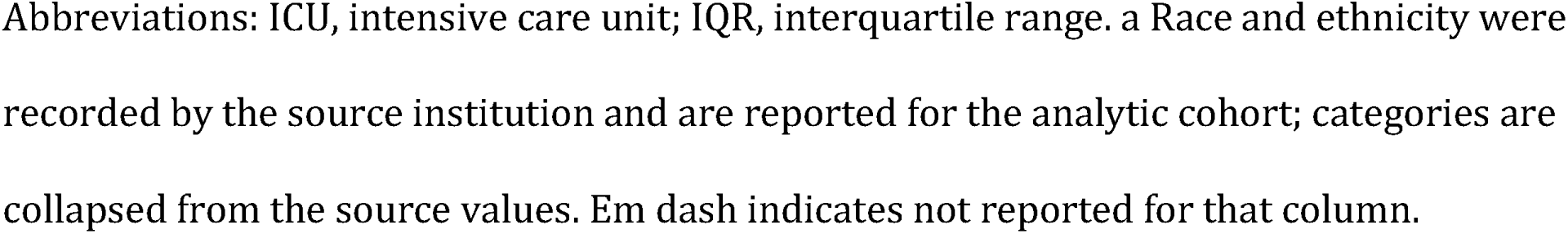
Characteristics of the ICU cohort.

| **Characteristic** | **Overall (N = 94,458)** | **Screened, analytic cohort (n = 72,944)** | **Never screened (n = 21,514)** |
| --- | --- | --- | --- |
| Age, median (IQR), y | 65 (53-76) | 65 (53-75) | 65 (52-77) |
| Women, No. (%) | 41,562 (44.0) | 32,095 (44.0) | 9,509 (44.2) |
| Primary language other than English, % | 10.0 | 10.0 | 9.7 |
| Race and ethnicity, %a |  |  |  |
| White | ... | 64.4 | ... |
| Black | ... | 11.1 | ... |
| Hispanic | ... | 3.8 | ... |
| Asian | ... | 3.2 | ... |
| Other or unknown | ... | 17.6 | ... |
| Any acute brain injury, % | 19.3 | 19.2 | 19.8 |
| Neuroscience ICU, % | 9.5 | 11.8 | 1.5 |
| In-hospital mortality, % | 12.0 | 12.1 | 11.6 |
| ICU length of stay, median, d | 2.0 | 2.1 | 1.7 |
| Delirium screens per stay, median (IQR) | ... | 5 (3-9) | 0 |
| Per-stay Unable-to-Assess fraction, median (IQR) | ... | 0 (0-0.20) | ... |

Abbreviations: ICU, intensive care unit; IQR, interquartile range. a Race and ethnicity were recorded by the source institution and are reported for the analytic cohort; categories are collapsed from the source values. Em dash indicates not reported for that column.

### More than one in five delirium screens recorded that the patient could not be examined

Of 610,632 screens, 130,455 (21.4%; 95% CI, 21.0%-21.8%) were Unable to Assess, exceeding the 119,052 (19.5%) scored Positive; 361,125 (59.1%) were Negative (Figure 1A). Unassessable results were present on 23.7% of assessed ICU-days, and 35.3% of stays carried at least one. The burden was concentrated (median per-stay UTA fraction, 0 [IQR, 0-0.20]) and stable in sensitivity analyses (21.9% for stays of at least 24 hours; 21.4% across 30-, 60-, and 120-minute and prior-only RASS-matching windows; eTable 11).

**Figure 1.**
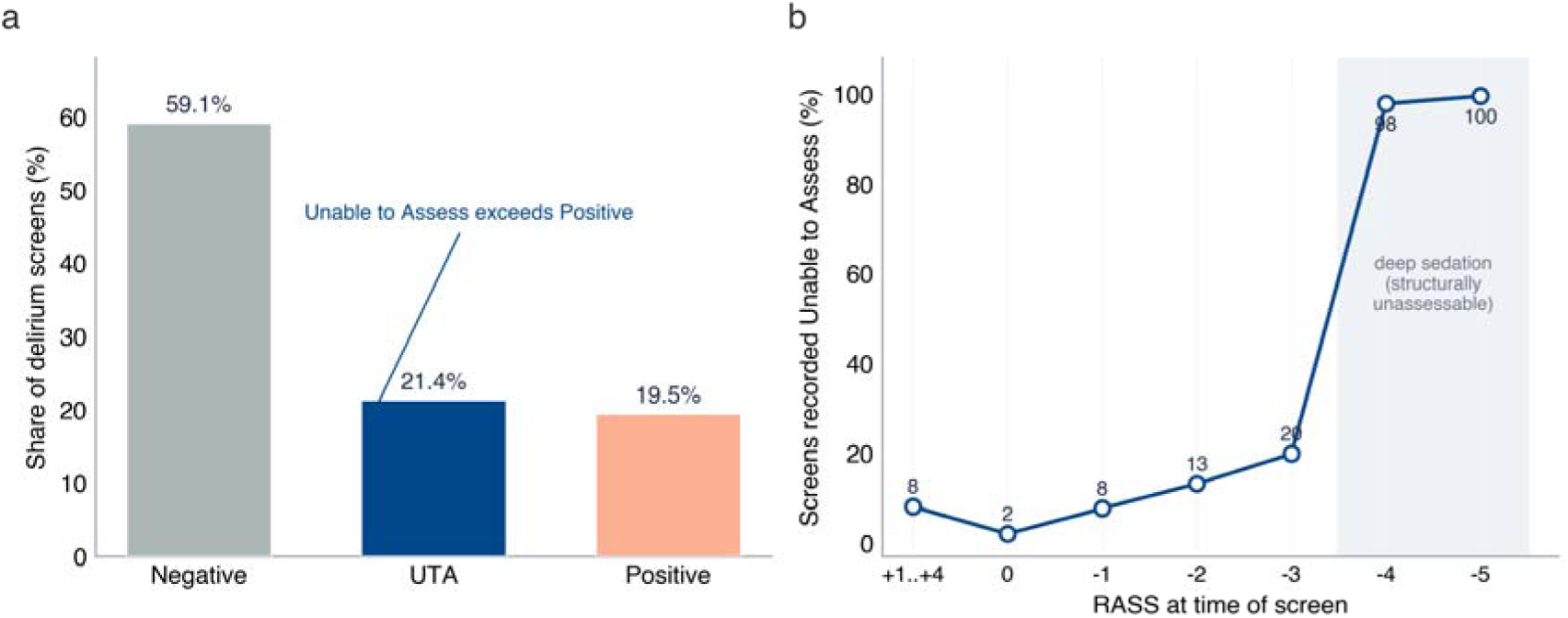
Burden and structural gradient of unassessable delirium screening. (A) Composition of 610,632 delirium screens. Unable to Assess (21.4%) exceeds Positive (19.5%). (B) Proportion of screens recorded as Unable to Assess by Richmond Agitation-Sedation Scale (RASS) at the time of the screen; the fraction rises from 2.0% at RASS 0 to 97.8% and 99.5% at RASS -4 and -5.

### Unable-to-Assess Documentation Tracks Arousal Almost Deterministically

The probability that a screen was UTA rose steeply as arousal fell (Figure 1B): 2.0% at RASS 0, 7.7% at RASS -1, 13.2% at -2, and 19.9% at -3, then jumped to 97.8% at RASS -4 and 99.5% at -5. Of all UTA screens, 101,388 (77.7%) occurred at RASS -4 or -5 and were structurally unassessable; 28,733 (22.0%) occurred in arousable patients (RASS -3 or higher), concentrated at the borderline RASS -3. Among clearly arousable screens (RASS -2 or higher), 4.6% were UTA.

In an adjusted model of UTA among arousable screens, the result was more likely with mechanical ventilation (OR, 3.43; 95% CI, 3.17-3.71), any acute brain injury (OR, 2.01; 95% CI, 1.87-2.16), and active sedation (OR, 1.43; 95% CI, 1.33-1.54), and less likely in neuroscience ICUs (OR, 0.35; 95% CI, 0.31-0.40); night shift and older age were also independently associated (eTable 12). The associations were similar or stronger among clearly arousable screens (RASS -2 or higher).

### Unassessable Screens Concentrate in the Patients Delirium Surveillance Most Targets

UTA documentation varied widely by acute brain-injury phenotype (Figure 2A). It was highest in status epilepticus (58.4%), anoxic or hypoxic-ischemic injury (56.3%), and cardiac arrest (43.5%), the phenotypes most often deeply sedated or comatose, and 25.0% to 27.2% across the stroke and trauma phenotypes. Screens from patients with any acute brain injury were unassessable 29.8% of the time versus 18.2% without (Cramér V, 0.125; P < .001).

**Figure 2.**
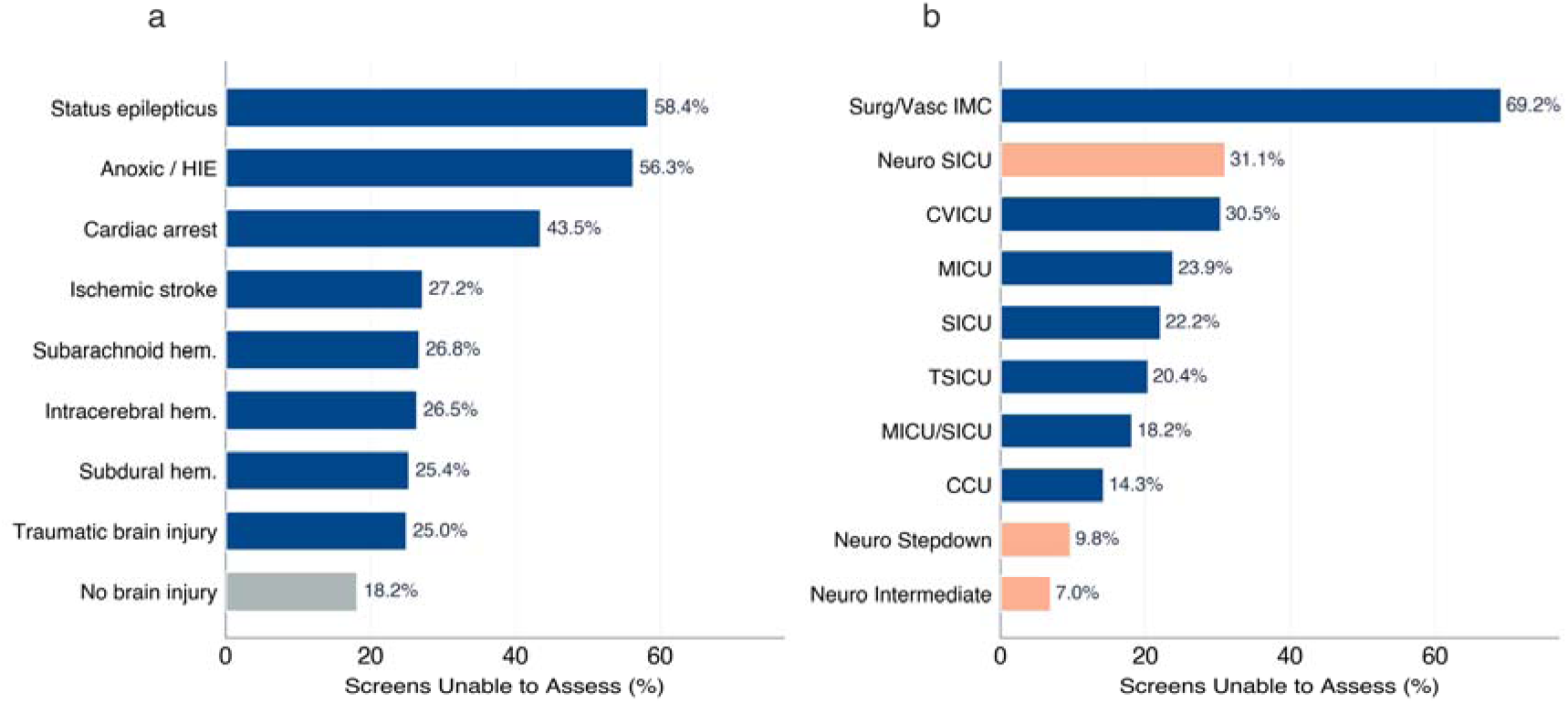
Concentration of unassessable screening. (A) Unable-to-Assess fraction by acute brain-injury phenotype and (B) by ICU type, ordered by magnitude.

Variation across ICU type was also marked (Figure 2B; Cramér V, 0.186; P < .001). The unassessable fraction was lowest in the neuroscience intermediate (7.0%) and stepdown (9.8%) units and highest in the medical (23.9%), cardiovascular (30.5%), and neuroscience surgical (31.1%) ICUs, the last reflecting postoperative sedation.

### The Racial and Ethnic Gap in Non-Assessment Among Arousable Screens Is Largely a Language Gap

Among arousable screens (RASS -3 or higher), UTA differed by race and ethnicity: relative to White patients (4.6%), the rate was higher in Asian (12.2%; relative risk [RR], 2.65; 95% CI, 2.53-2.77), Hispanic (10.5%; RR, 2.28; 95% CI, 2.18-2.39), and Black patients (6.4%; RR, 1.38; 95% CI, 1.34-1.43). The gradient corresponded to language: UTA among arousable screens was 4.5% for English-speaking and 16.2% for non-English-speaking patients, a gap present across ICU type and shift (eTable 13). After adjustment for non-English language together with arousal, sedation, ventilation, brain injury, age, and sex, the racial and ethnic ORs were substantially attenuated (Asian, 1.30 [95% CI, 1.12-1.52]; Hispanic, 1.17 [95% CI, 1.01-1.36]; Black, 1.13 [95% CI, 1.01-1.26]), while non-English language retained an OR of 3.74 (95% CI, 3.43-4.08; E-value, 6.95; eMethods), rising to 5.01 (95% CI, 4.63-5.43) among clearly arousable screens (RASS -2 or higher) (Figure 3). The non-English association was essentially unchanged after adding ICU-type and admission-era fixed effects (OR, 3.94; 95% CI, 3.61-4.30; eTable 16). Adding primary payer to the same model left the non-English association essentially unchanged (OR, 3.65; 95% CI, 3.34-3.99) and revealed a smaller independent insurance gradient: relative to private insurance, unassessment among arousable screens was modestly more frequent for other or unknown insurance (adjusted OR, 1.40; 95% CI, 1.18-1.66) and Medicaid (adjusted OR, 1.13; 95% CI, 1.01-1.25), but not Medicare (adjusted OR, 1.08; 95% CI, 0.98-1.19); absolute UTA rates rose from 4.1% under private insurance to 6.0% (Medicare), 6.7% (Medicaid), and 7.5% (other or unknown), and only the other-or-unknown contrast survived correction across the full equity family (eTable 20). Language, not insurance, remained the dominant and most robust correlate of avoidable non-assessment.

**Figure 3.**
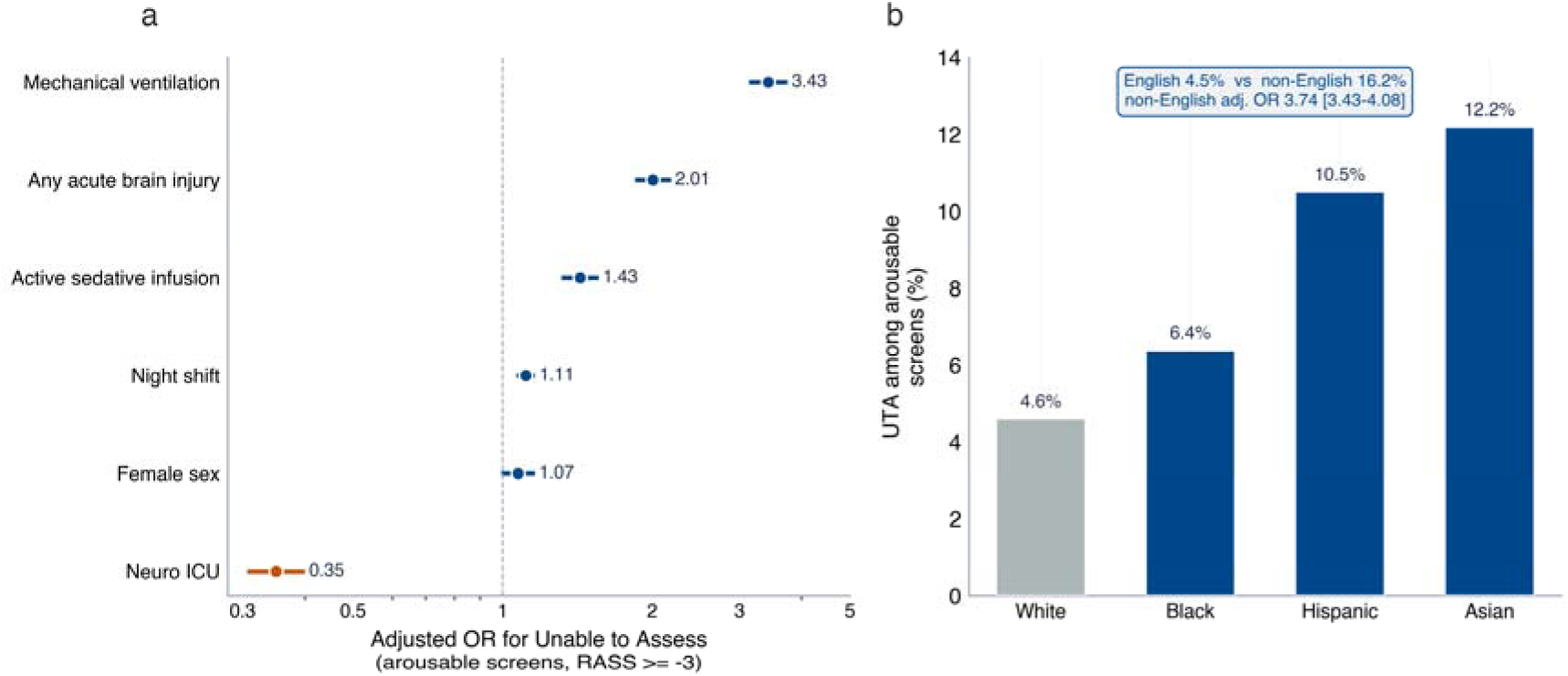
Non-assessment among arousable screens, and language. (A) Adjusted odds ratios for Unable to Assess among arousable screens (RASS -3 or higher). (B) Unable-to-Assess rate among arousable screens by patient race and ethnicity, with the English versus non-English contrast and the language-adjusted odds ratio annotated. The non-English association strengthens for potentially avoidable non-assessment (clearly arousable screens, RASS -2 or higher; eMethods).

### How Unable-to-Assess Screens Are Handled Changes What the Data Appear to Show

Building the stay-level delirium label three ways from the same patients changed downstream estimates, modestly for some quantities and substantially for others (Figure 4). Crude delirium prevalence shifted only slightly, from 32.1% under complete-case handling to 30.8% when UTA was recoded as Negative and 31.7% under missingness-aware handling (2,544 stays indeterminate), and a simple delirium-prediction model’s out-of-fold AUROC shifted from 0.737 to 0.719. The delirium-to-in-hospital-mortality association was far more sensitive to the choice. In the primary model, adjusted for age, sex, the Charlson Comorbidity Index, and acute brain injury, the OR was 4.12 (95% CI, 3.88-4.36) under complete-case handling and 3.77 (95% CI, 3.56-3.99) under missingness-aware handling but fell to 2.16 (95% CI, 2.06-2.27) when UTA was recoded as Negative, a near-halving driven by reassigning deeply sedated, high-mortality patients to the non-delirium column. Complete-case handling, in turn, silently dropped 3,065 all-UTA stays whose mortality far exceeded that of retained stays (65.2% vs 9.8%; eTable 17). The pattern held in a secondary model that additionally adjusted for mechanical ventilation, vasopressor use, and log length of stay, although those mediators attenuated every estimate (complete-case 3.13 [95% CI, 2.92-3.35], missingness-aware 2.82 [2.64-3.02], recode-as-negative 1.66 [1.56-1.77]). A quantitative bias analysis showed that the recode-as-negative estimate is a lower bound that rises with the assumed delirium prevalence among unassessable screens (eTable 18).

**Figure 4.**
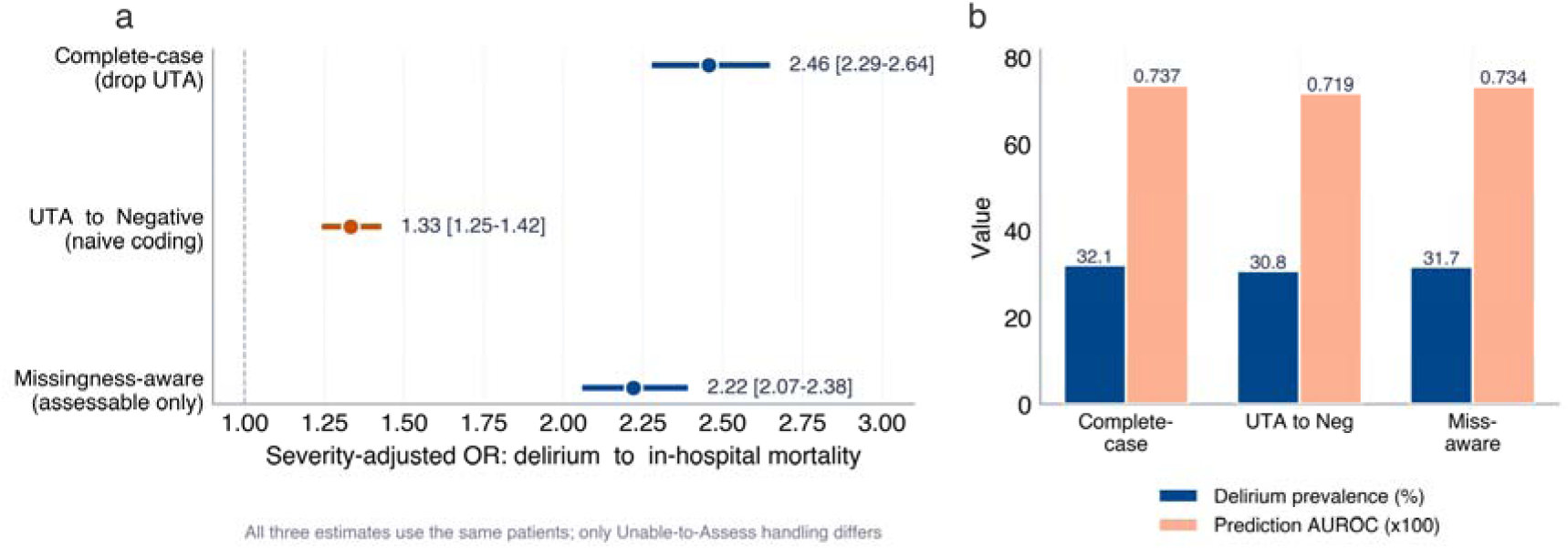
Downstream analytic consequences of handling choices. (A) Delirium-to-in-hospital-mortality odds ratio under three handling strategies for the same patients, adjusted for age, sex, the Charlson Comorbidity Index, and acute brain injury (primary model; a secondary model additionally adjusted for mechanical ventilation, vasopressor use, and log length of stay); the contrast is an analytic sensitivity to handling, not a causal estimate. (B) Delirium prevalence and prediction-model AUROC under each strategy.

### Unassessability Is Recoverable From the Observed Clinical State

A model predicting the UTA label from the patient’s structural state alone discriminated almost perfectly (gradient boosting AUROC, 0.951 [95% CI, 0.950-0.951]; logistic regression, 0.930; Brier score, 0.044) (eFigure 1). Because the features are close to definitional, this is evidence that unassessability is recoverable from the observed clinical state and is therefore informative missingness. All 11 tests in the primary inferential family remained significant after Benjamini-Hochberg correction.

### The Arousal Mechanism Replicates in an Independent Multicenter Cohort

In the eICU Collaborative Research Database (208 U.S. hospitals; eMethods S1.10), delirium screening was documented at 56 hospitals. Among 23,968 stays that both screened and recorded RASS, a scored delirium result was present in 24.2% of arousable stay-hours (RASS -3 or higher) but only 9.7% of deeply sedated ones (RASS -4 or -5), an odds of 1.24 per one-point increase in RASS (95% CI, 1.24 to 1.25; eTable 21). eICU records no UTA token, so unassessability appears as absence of a scored screen; the arousal mechanism therefore generalizes across two opposite documentation conventions.

## Discussion

In a large critical-care database, more than one in five recorded delirium screens stated that the patient could not be examined, a larger share than were scored positive. Three quarters fell on patients in deep sedation or coma, where the instrument does not apply; the remainder fell on arousable patients and was concentrated at the borderline RASS -3, where assessability is genuinely uncertain. The potentially avoidable share (RASS -2 or higher; 4.6% of such screens) was smaller but systematically patterned, falling on the ventilated, the brain-injured, and patients who did not speak English.

"Unable to Assess" is a faithful record of a real clinical situation, not a documentation error to clean away. Its near-deterministic relationship with arousal, recoverable at an AUROC above 0.93, marks it as informative missingness.^14^ Recoding nearly halved the apparent delirium-death association by assigning deeply sedated, high-mortality patients to the negative column;^2,15^ complete-case handling avoids that but removes the sickest patients from the denominator. Neither default is harmless, and the choice is rarely reported.

These findings extend a single-center literature that treated unassessable documentation as a bedside quality problem.^7^ We recast it as a measurement problem for secondary research. Contemporary delirium studies built on the same database exclude UTA patients as a complete-case step or define delirium only from positive screens, so unassessable windows can silently become non-delirium (eTable 15).^17–20^ A systematic review found delirium-model missing-data handling frequently poorly documented or methodologically unsound.^21^

The concentration in acute brain injury has two sources this study cannot separate: depressed arousal (from sedation or injury) and overlap between the delirium construct and acute neurologic deficits, in which a patient may be arousable yet untestable by a tool requiring verbal attention tasks, as with aphasia after dominant-hemisphere stroke. Both leave delirium status unknown in the patients whose encephalopathy most warrants surveillance.

The CAM-ICU requires verbal interaction, so a tool that cannot be administered across a language barrier leaves a surveillance gap for patients who do not speak the language of care.^16^ That the racial and ethnic differences were substantially attenuated once language was included, while non-English language retained a threefold to fivefold association across ICU type, shift, and admission era, points to an addressable target, interpreter-supported or language-concordant assessment, rather than a property of any patient group. Lower non-assessment in neuroscience intermediate and stepdown units suggests the gap is also responsive to unit culture and staffing.

This study has limitations. It draws on a single US academic center; absolute frequencies depend on local screening policy, documentation conventions, staffing, sedation practice, and interpreter workflows, although the arousal mechanism was externally validated in eICU. Screening practice changed over time, which colors the never-screened fraction. The borderline RASS -3 stratum is of uncertain assessability; even at RASS -2 or higher, the data cannot confirm an individual screen was truly completable because interpreter availability, aphasia, sensory impairment, staffing, and cooperation are not recorded. MIMIC-IV has no structured neurologic outcome scale, and the delirium-mortality estimates illustrate sensitivity to handling, not causality. The UTA label reflects nurse documentation and was not audited against a gold-standard examination, though it was reproduced by independent CAM-ICU feature items.

Delirium datasets should preserve "Unable to Assess" as a distinct state; every delirium study should report how it was handled; and prediction-model reports should disclose excluded unassessable screens. The unassessable fraction can serve as its own quality metric, stratified by arousal and language, and its concentration in arousable non-English-speaking patients is a measurable equity target.

Where delirium surveillance is universal policy, the most common recorded result after "negative" was not "positive" but "could not be assessed." Treating that result as if it carried no information misstates both how much delirium is seen and how strongly it tracks death. The unexaminable patient is not a gap in the data to be discarded but an informative state that secondary analyses should preserve and report.

## Supporting information

Full_appendix

## Data Availability

All data produced are available online at https://github.com/Alon-Gorenshtein/study_delirium_unscreenable

