## Supplementary material for "Unscreenable: The Burden, Structure, and Analytic Consequences of "Unable to Assess" Delirium Documentation in the Intensive Care Unit": Full_appendix

This Supplementary Information accompanies the main manuscript. It contains the full measurement and statistical methods (eMethods), supplementary tables (eTable 1 to eTable 21), and one supplementary figure (eFigure 1). All analyses used the Medical Information Mart for Intensive Care IV (MIMIC-IV) version 3.1 under credentialed PhysioNet access.

**Contents**

- S1. Supplementary Methods (eMethods)
- S2. Supplementary Tables (eTable 1 to eTable 21)
- S3. Supplementary Figures (eFigure 1)
- S4. Code and Data Availability

### S1. Supplementary Methods

#### S1.1 Data source and cohort

MIMIC-IV version 3.1 contains de-identified records for adult patients admitted to the intensive care units (ICUs) of a single academic medical center between 2008 and 2019. All adult ICU stays (94,458 stays) were eligible. The analytic cohort comprised the 72,944 stays with at least one charted delirium screen. Stays with no delirium screen (21,514) were retained as a separate denominator to quantify the never-screened fraction.

#### S1.2 Delirium screen variable

Delirium screens were taken from the charted item ”Delirium assessment” (itemid 228332), which records each screen as one of three states: Negative, Positive, or Unable to Assess (UTA). This item is the nurse-recorded result of the Confusion Assessment Method for the ICU (CAM-ICU) workflow. The three states are mutually exclusive and exhaustive; no free-text parsing was required. The eight CAM-ICU feature items (itemids 228300 to 228303 and 228334 to 228337), which separately record an ”Unable to Assess” response on individual features, were extracted for the construct-validity analysis in eTable 9.

#### S1.3 Arousal indexing and the structural versus potentially avoidable split

Arousal at each screen was indexed with the Richmond Agitation-Sedation Scale (RASS, itemid 228096). RASS values are stored as free text with a leading signed integer and a descriptor (for example, "-4 Deep sedation"), occasionally carrying a stray leading quotation mark from the source export; the leading signed integer was parsed with the regular expression `^[\s"']*([+-]?\d+)` and retained when in the valid range -5 to +4. Each screen was matched to the nearest RASS reading within the same ICU stay within a 2-hour window using a nearest-neighbor as-of join; a RASS value was matched for 99.8% of screens.

Each UTA screen was classified by the matched RASS as:

- **Structurally unassessable** (RASS -4 or -5): the patient was in deep sedation or unarousable, a state in which the CAM-ICU is not applicable by protocol.
- **Arousable** (RASS -3 or higher): the patient was arousable enough that the CAM-ICU was potentially completable. Because RASS -3 is borderline, it is reported separately; potentially avoidable non-assessment was defined conservatively as UTA at RASS -2 or higher (eTable 11 to eTable 13).

#### S1.4 Sedation, neuromuscular blockade, and ventilation

Active drug exposure at the time of each screen was derived from timestamped infusion intervals (input events). Sedatives comprised propofol (itemid 222168), midazolam (221668), dexmedetomidine (225150, 229420), fentanyl (221744, 225942, 225972), ketamine (221712), lorazepam (221385), and pentobarbital (225156). Neuromuscular blockers comprised cisatracurium (221555), vecuronium (222062), and rocuronium (229233). Mechanical ventilation was derived from procedure-event spans (invasive ventilation 225792, intubation 224385). A screen was flagged as coincident with an exposure when its chart time fell inside a documented interval for that exposure.

#### S1.5 Acute brain-injury phenotypes

Phenotypes were defined by version-aware International Classification of Diseases, Ninth and Tenth Revision (ICD-9 and ICD-10) codes in any diagnosis position (eTable 2). The composite "any acute brain injury" flag combined ischemic stroke, intracerebral hemorrhage, subarachnoid hemorrhage, subdural hemorrhage, traumatic brain injury, anoxic or hypoxic-ischemic injury, cardiac arrest, and status epilepticus.

#### S1.6 Statistical analysis

Proportions are reported with Wilson 95% confidence intervals. Screen-level proportions also carry patient-clustered bootstrap confidence intervals (1000 resamples, resampling patients with replacement). Categorical associations between the UTA indicator and phenotype or ICU type were tested with the chi-square test and summarized with the Cramer V effect size. Adjusted associations were estimated with logistic regression fitted by generalized estimating equations with cluster-robust variance by patient, reporting odds ratios with 95% confidence intervals.

Two models of the UTA indicator were fitted. The first used all screens with a matched RASS and entered deep sedation (RASS -4 or lower) and moderate sedation (RASS -3) as indicators; because deep sedation is nearly deterministic of UTA, its odds ratio is extremely large and is reported for transparency only (eTable 4). The second, the prespecified analytic model, was restricted to arousable screens (RASS -3 or higher) and modeled UTA among arousable screens on patient and process covariates, where no separation occurs. Mechanical ventilation is a mediator of the sedation-to-UTA pathway and was reported but not used to adjust the sedation estimate.

The equity analysis modeled UTA among arousable screens (RASS -3 or higher) on race and ethnicity, first unadjusted (relative risk with the Katz log method, eTable 6) and then adjusted for non-English primary language together with arousal, sedation, ventilation, brain injury, age, and sex, to test whether language attenuated racial and ethnic differences. For the adjusted non-English-language association, the E-value for unmeasured confounding was computed by the VanderWeele and Ding formula for an odds ratio above 1 (E = OR + sqrt[OR x (OR - 1)]), applied to the point estimate and to the lower confidence limit.

The downstream-bias analysis (eTable 7) built a stay-level delirium label three ways from the same cohort: complete-case (screens recorded as UTA dropped, and a stay labeled positive if any retained screen was positive), UTA recoded as Negative, and missingness-aware (positive among assessable screens only, with stays that had no assessable screen reported separately as indeterminate). Each strategy estimates a distinct quantity: complete-case (delirium among patients with a scorable screen), UTA-as-Negative (apparent stay-level delirium), and missingness-aware (delirium among assessable screens). Under each we computed delirium prevalence, the delirium-to-in-hospital-mortality odds ratio (cluster-robust by patient), and the out-of-fold area under the receiver operating characteristic curve (AUROC) of a delirium-prediction model. The mortality odds ratio is reported from a baseline-adjusted model (age, sex, the Charlson Comorbidity Index, acute brain injury) as the primary analysis, a secondary model that additionally adjusts for mechanical ventilation, vasopressor use, and log length of stay (mediators on the delirium-to-death path), and an age- and sex-only model for comparison (eTable 14); all are analytic sensitivities to handling, not causal estimates. Robustness of the UTA fraction and the potentially avoidable split to the RASS-matching window (30, 60, 120 minutes, and prior-RASS-only) is in eTable 11. The prediction model used age, sex, ever-ventilated, ever-sedated, neuroscience-ICU, and any-brain-injury features in a standardized-feature logistic-regression pipeline with 5-fold stratified cross-validation.

P values across the primary inferential family of 11 tests (eTable 8) were adjusted with the

Benjamini-Hochberg false-discovery-rate procedure at a threshold of 0.05, two-sided. Analyses used Python 3.9 with pandas 2.3, NumPy 2.0, SciPy 1.13, statsmodels 0.14.6, and scikit-learn 1.6.1.

#### S1.7 Unscreenability probe (model card)

The probe quantifies how systematically unassessability is determined by the patient's structural state. It is a measurement instrument for the missingness mechanism, not a clinical prediction tool, and it was not externally validated.

- **Task**: predict whether a single delirium screen is recorded as UTA.
- **Population**: all screens with a matched RASS (609,139 screens).
- **Features**: RASS, active sedation, active neuromuscular blockade, mechanical ventilation, hour of day, neuroscience-ICU indicator, and the eight brain-injury phenotype flags.
- **Estimators**: L2-regularized logistic regression in a standardized-feature pipeline, and a histogram gradient-boosting classifier (scikit-learn 1.6.1, default hyperparameters).
- **Validation**: 5-fold cross-validation grouped by patient (no patient appears in both training and test folds). Out-of-fold predictions were pooled for evaluation.
- **Metrics**: AUROC and Brier score, with 95% confidence intervals from a 1000-sample percentile bootstrap; the two estimators were compared with a paired bootstrap of out-of-fold predictions.
- **Result**: logistic AUROC 0.930 (95% CI, 0.929 to 0.931); gradient-boosting AUROC 0.951 (95% CI, 0.950 to 0.951); Brier 0.044; paired difference +0.021 favoring gradient boosting.

#### S1.8 Construct-validity sensitivity analysis

The "Unable to Assess" state of the summary item (228332) was cross-checked against the independent CAM-ICU feature items (228334 to 228337 and 228300), which separately record an "Unable to Assess" response. Each of these items carried a substantial UTA fraction (eTable 9), confirming that unassessability is an explicit, recurring recorded state rather than an artifact of a single field.

#### S1.9 Revision sensitivity analyses

Four additional analyses were added in revision (eTable 16 to eTable 19); each is a sensitivity analysis, not a primary estimate.

- **Language model with fixed effects (eTable 16).** The Figure 3 adjusted model of UTA among arousable screens was refitted with ICU-type and admission-era (anchor-year-group) fixed effects added, to test whether the non-English-language association reflected unit mix or calendar-era documentation. Estimation was by cluster-robust logistic regression.
- **Stays removed under complete-case handling (eTable 17).** Stays scored only as UTA (dropped by complete-case handling) and stays with no assessable screen (reported as indeterminate by missingness-aware handling) were compared with retained stays on mortality, organ support, brain injury, language, unit, length of stay, and the proportion of their screens at deep sedation (RASS -4 or lower).
- **Quantitative bias analysis (eTable 18).** Because the true delirium status of an unassessable screen is unknown, the UTA-as-negative strategy implicitly assumes it is zero. We varied the assumed delirium prevalence among UTA screens from 0% to 100% and, at each value, randomly reassigned that fraction of currently non-positive stays carrying a UTA screen to positive (30 replicates, fixed seed), recomputing stay-level prevalence and the baseline-adjusted mortality odds ratio.
- **Inverse-probability weighting and augmentation (eTable 19).** Restricted to arousable screens, the assessability propensity P(scored | observed state) was estimated by cross-fitted logistic regression, and inverse-probability-weighted (Hajek) and augmented (doubly robust)

estimators were formed for the delirium-positive prevalence that would obtain had every arousable screen been scorable, under missingness at random given observed state. Weights were truncated at probabilities of 0.02 and 0.98. The estimator is defensible only within arousable states and is reported as a bounding sensitivity, not the true prevalence, because extending missingness at random into structurally unassessable (RASS -4 or lower) screens is not warranted.

The literature characterization in eTable 15 was assembled by targeted search of delirium studies using MIMIC-IV and related databases; it is illustrative rather than a systematic review, and each study's handling of unassessable screens was read from its published methods.

#### S1.10 External validation of the arousal mechanism (eICU)

To test whether the arousal dependence of assessability generalizes beyond this center, we replicated the mechanism in the eICU Collaborative Research Database (eICU-CRD) version 2.0, a multicenter database of more than 200,000 intensive care unit stays at 208 U.S. hospitals (2014 to 2015), accessed through PhysioNet. eICU records no Unable-to-Assess token; a delirium screen (nurseCharting cell label ”Delirium Scale/Score”, value name ”Delirium Score”) appears only when it was scored Yes, No, or as a numeric value, so unassessability is expressed as the absence of a screen rather than a labeled state. The Richmond Agitation-Sedation Scale was taken from the sedation flowsheet (Sedation Scale/Score/Goal with Sedation Scale = ”RASS”, value in Sedation Score), because the dedicated RASS field and the delirium screen were charted by disjoint hospitals. Among stays that both screened and recorded a RASS, we measured (1) the RASS recorded nearest each scored screen (within 120 minutes) and (2) the probability that a scored result was present in a stay-hour given the deepest RASS that hour, with a logistic odds ratio per one-point increase in RASS. Because delirium screening was documented at only a minority of hospitals, we make no claim about the population unassessable fraction; eICU has no language field, so the demographic patterning was not externally tested.

### S2. Supplementary Tables

**eTable 1. Variable definitions and source items.** All items are from the MIMIC-IV ICU module unless noted.

| Variable | Source | Item or field |
|---|---|---|
| Delirium screen result | chartevents | itemid 228332 (Negative / Positive / Unable to Assess) |
| CAM-ICU feature items | chartevents | itemids 228300-228303, 228334-228337 |
| Richmond Agitation-Sedation Scale | chartevents | itemid 228096 |
| Sedative infusions | inputevents | propofol 222168, midazolam 221668, dexmedetomidine 225150 / 229420, fentanyl 221744 / 225942 / 225972, ketamine 221712, lorazepam 221385, pentobarbital 225156 |
| Neuromuscular blockers | inputevents | cisatracurium 221555, vecuronium 222062, rocuronium 229233 |
| Mechanical ventilation | procedureevents | invasive ventilation 225792, intubation 224385 |
| ICU type | icustays | first_careunit |
| Demographics, language, outcomes | admissions, patients | race, insurance, language, anchor_age, gender, hospital_expire_flag, length of stay |

**eTable 2. Acute brain-injury phenotype definitions.** Any-position ICD-9 and ICD-10 code

prefixes.

| Phenotype | ICD-9 prefix | ICD-10 prefix |
|---|---|---|
| Ischemic stroke | 433, 434 | I63 |
| Intracerebral hemorrhage | 431 | I61 |
| Subarachnoid hemorrhage | 430 | I60 |
| Subdural hemorrhage | 432 | I62 |
| Traumatic brain injury | 800-804, 850-854 | S06 |
| Anoxic or hypoxic-ischemic injury | 348.1, 348.5 | G93.1, G93.82 |
| Cardiac arrest | 427.5 | I46 |
| Status epilepticus | 345.3 | G41 |

**eTable 3. Unable-to-Assess fraction by Richmond Agitation-Sedation Scale (RASS) at the time of the screen.** Screens with a matched RASS.

| RASS | Screens, No. | Unable to Assess, % |
|---|---:|---:|
| +1 to +4 | 55,342 | 8.1 |
| 0 | 294,463 | 2.0 |
| -1 | 83,624 | 7.7 |
| -2 | 38,365 | 13.2 |
| -3 | 34,573 | 19.9 |
| -4 | 51,847 | 97.8 |
| -5 | 50,925 | 99.5 |

**eTable 4. Adjusted odds ratios for an Unable-to-Assess result.** Cluster-robust logistic regression. The arousable-only model (RASS -3 or higher) is the prespecified analytic model. The full model is shown for transparency; the deep-sedation term reflects near-deterministic separation

and should not be interpreted as a calibrated effect.

| Covariate | Arousable model OR (95% CI) | Full model OR (95% CI) |
|---|---|---|
| Mechanical ventilation | 3.43 (3.17, 3.71) | 2.83 (2.60, 3.07) |
| Any acute brain injury | 2.01 (1.87, 2.16) | 1.82 (1.70, 1.96) |
| Active sedative infusion | 1.43 (1.33, 1.54) | 1.22 (1.13, 1.31) |
| Night shift | 1.11 (1.08, 1.15) | 1.10 (1.07, 1.14) |
| Female sex | 1.07 (1.00, 1.15) | not entered |
| Neuroscience ICU | 0.35 (0.31, 0.40) | 0.36 (0.33, 0.40) |
| Age, per year | 1.02 (1.01, 1.02) | not entered |
| Moderate sedation (RASS -3) | not entered | 2.57 (2.43, 2.72) |
| Deep sedation (RASS -4 or lower) | not entered | 1030 (955, 1112) |

**eTable 5. Unable-to-Assess fraction by acute brain-injury phenotype and by ICU type.** ICU types with at least 2000 screens are shown.

| Group | Unable to Assess, % | Screens, No. |
|---|---:|---:|
| **Phenotype** | | |
| Status epilepticus | 58.4 | 5,348 |
| Anoxic or hypoxic-ischemic injury | 56.3 | 29,610 |
| Cardiac arrest | 43.5 | 30,501 |
| Ischemic stroke | 27.2 | 58,470 |
| Subarachnoid hemorrhage | 26.8 | 21,855 |
| Intracerebral hemorrhage | 26.5 | 38,196 |
| Subdural hemorrhage | 25.4 | 10,809 |
| Traumatic brain injury | 25.0 | 29,762 |
| Any acute brain injury | 29.8 | 165,250 |

| Group | Unable to Assess, % | Screens, No. |
|---|---|---|
| No acute brain injury | 18.2 | 445,382 |
| **ICU type** | | |
| Neuroscience surgical ICU | 31.1 | 14,286 |
| Cardiovascular ICU | 30.5 | 81,758 |
| Medical ICU | 23.9 | 167,990 |
| Surgical ICU | 22.2 | 82,536 |
| Trauma surgical ICU | 20.4 | 57,334 |
| Medical-surgical ICU | 18.2 | 80,342 |
| Coronary care unit | 14.3 | 54,886 |
| Neuroscience stepdown | 9.8 | 12,984 |
| Neuroscience intermediate | 7.0 | 53,012 |

**eTable 6. Unable-to-Assess among arousable screens (RASS -3 or higher) by race, ethnicity, insurance, and language.** Relative risks are unadjusted (reference, White or Private). Adjusted odds ratios are from the model that includes non-English language, arousal, sedation, ventilation, brain injury, age, and sex. Potentially avoidable non-assessment (the stricter RASS -2 or higher definition) is in eTable 12.

| Group | UTA among arousable, % | Relative risk (95% CI) | Adjusted OR (95% CI) |
|---|---|---|---|
| White | 4.6 | 1 [Reference] | 1 [Reference] |
| Black | 6.4 | 1.38 (1.34, 1.43) | 1.13 (1.01, 1.26) |
| Hispanic | 10.5 | 2.28 (2.18, 2.39) | 1.17 (1.01, 1.36) |
| Asian | 12.2 | 2.65 (2.53, 2.77) | 1.30 (1.12, 1.52) |
| English language | 4.5 | 1 [Reference] | 1 [Reference] |
| Non-English language | 16.2 | ... | 3.74 (3.43, 4.08) |

| Group | UTA among arousable, % | Relative risk (95% CI) | Adjusted OR (95% CI) |
|---|---|---|---|
| Private insurance | 4.1 | 1 [Reference] | ... |
| Medicare | 6.0 | 1.46 | ... |
| Medicaid | 6.7 | 1.64 | ... |

**eTable 7. Downstream analytic consequences of three Unable-to-Assess handling strategies.** Same cohort; only the handling of UTA screens differs. The mortality odds ratio is from the primary baseline-adjusted model (age, sex, the Charlson Comorbidity Index, acute brain injury); the mediator-adjusted and age- and sex-only values are in eTable 14. All are analytic sensitivities to handling, not causal estimates.

| Strategy | Stays, No. | Delirium prevalence, % (95% CI) | Baseline-adjusted mortality OR (95% CI) | Prediction AUROC |
|---|---|---|---|---|
| Complete-case (drop UTA) | 69,879 | 32.1 (31.8, 32.5) | 4.12 (3.88, 4.36) | 0.737 |
| UTA recoded as Negative | 72,944 | 30.8 (30.5, 31.1) | 2.16 (2.06, 2.27) | 0.719 |
| Missingness-aware | 70,400 | 31.7 (31.4, 32.1) | 3.77 (3.56, 3.99) | 0.734 |

Under the missingness-aware strategy, 2544 stays had no assessable screen and were reported as indeterminate rather than scored.

**eTable 8. Primary inferential family with Benjamini-Hochberg correction.** All 11 tests remained significant at a false-discovery-rate threshold of 0.05. The delirium-mortality rows report the baseline-adjusted estimates used in the main text and eTable 7; the mediator-adjusted and age- and sex-only estimates (eTable 14) show the same direction and significance.

| Test | Effect | 95% CI | Adjusted P |
|---|---|---|---|
| UTA by any brain injury (Cramer V) | 0.125 | ... | <0.001 |
| UTA by ICU type (Cramer V) | 0.186 | ... | <0.001 |
| UTA among arousable screens, mechanical ventilation (OR) | 3.43 | 3.17, 3.71 | <0.001 |
| UTA among arousable screens, any brain injury (OR) | 2.01 | 1.87, 2.16 | <0.001 |
| UTA among arousable screens, active sedation (OR) | 1.43 | 1.33, 1.54 | <0.001 |
| UTA among arousable screens, neuroscience ICU (OR) | 0.35 | 0.31, 0.40 | <0.001 |
| UTA among arousable screens, night shift (OR) | 1.11 | 1.08, 1.15 | <0.001 |
| UTA among arousable screens, non-English language (OR) | 3.74 | 3.43, 4.08 | <0.001 |
| Delirium-mortality OR, complete-case (baseline-adjusted) | 4.12 | 3.88, 4.36 | <0.001 |
| Delirium-mortality OR, UTA as Negative (baseline-adjusted) | 2.16 | 2.06, 2.27 | <0.001 |
| Delirium-mortality OR, missingness-aware (baseline-adjusted) | 3.77 | 3.56, 3.99 | <0.001 |

**eTable 9. Construct-validity check: Unable-to-Assess fraction in the independent CAM-ICU feature items.** Each CAM-ICU feature item separately records an ”Unable to Assess” response.

| CAM-ICU feature item | itemid | Stays, No. | Unable to Assess, % |
|---|---|---|---|
| Mental-status change | 228300 | 1,665 | 21.5 |
| Disorganized thinking | 228335 | 2,265 | 19.2 |
| Inattention | 228336 | 9,047 | 10.1 |
| Mental-status change | 228337 | 23,703 | 8.8 |
| Summary delirium assessment (main analysis) | 228332 | 72,944 | 21.4 |

#### S2.1 Revision sensitivity analyses

**eTable 10. Screening rate and Unable-to-Assess burden by era and ICU length of stay.**

Screened fraction is the proportion of ICU stays with at least one delirium screen; the UTA column is the mean per-stay UTA fraction among screened stays.

| Stratum | Screened, % | Mean per-stay UTA, % |
|---|---|---|
| **Admission era** | | |
| 2008-2010 | 43 | 12.4 |
| 2011-2013 | 84 | 14.8 |
| 2014-2016 | 98 | 14.5 |
| 2017-2019 | 97 | 14.1 |
| **ICU length of stay** | | |
| <1 day | 69 | 11.1 |
| 1-2 days | 77 | 13.4 |
| 2-5 days | 80 | 13.6 |
| >5 days | 82 | 22.5 |

**eTable 11. Robustness of the Unable-to-Assess fraction to the RASS-matching window.**

Each row re-matches every screen to a RASS reading under a different rule.

| Matching rule | Matched, % | UTA, % | UTA among arousable, % |
|---|---|---|---|
| Nearest within 30 min | 99.3 | 21.4 | 5.63 |
| Nearest within 60 min | 99.4 | 21.4 | 5.64 |
| Nearest within 120 min (main) | 99.8 | 21.4 | 5.67 |
| Prior RASS only, within 120 min | 99.4 | 21.4 | 5.64 |

**eTable 12. Adjusted odds ratios for Unable to Assess restricted to clearly arousable screens (RASS -2 or higher).** Cluster-robust logistic regression; the stricter arousability definition removes the borderline RASS -3 stratum.

| Covariate | OR (95% CI) |
|---|---|
| Non-English primary language | 5.01 (4.63, 5.43) |
| Mechanical ventilation | 3.01 (2.75, 3.30) |
| Any acute brain injury | 2.00 (1.85, 2.17) |
| Active sedative infusion | 1.43 (1.31, 1.57) |

**eTable 13. Unable-to-Assess among arousable screens (RASS -3 or higher), by language within ICU type and shift.**

| Stratum | English, % | Non-English, % |
|---|---|---|
| Neuroscience ICU | 1.1 | 10.8 |
| General (non-neuroscience) ICU | 5.0 | 17.3 |
| Day shift | 4.2 | 16.3 |
| Night shift | 4.8 | 16.1 |

**eTable 14. Delirium-to-in-hospital-mortality odds ratio under each handling strategy, by adjustment set.** The primary baseline model adjusts for age, sex, the Charlson Comorbidity Index, and acute brain injury; the mediator-adjusted model additionally adds mechanical ventilation, vasopressor use, and log length of stay (mediators on the delirium-to-death path); the minimal model adjusts for age and sex only. All are analytic sensitivities to handling, not causal estimates.

| Handling strategy | Minimal (age, sex) OR (95% CI) | Primary baseline OR (95% CI) | Mediator-adjusted OR (95% CI) |
|---|---|---|---|
| Complete-case (drop UTA) | 4.57 (4.32, 4.83) | 4.12 (3.88, 4.36) | 3.13 (2.92, 3.35) |
| UTA recoded as Negative | 2.54 (2.42, 2.66) | 2.16 (2.06, 2.27) | 1.66 (1.56, 1.77) |
| Missingness-aware | 4.20 (3.97, 4.43) | 3.77 (3.56, 3.99) | 2.82 (2.64, 3.02) |

#### S2.2 Revision sensitivity analyses, round 2

**eTable 15. Handling of ”Unable to Assess” in contemporary ICU delirium studies using MIMIC and related databases.** Studies are shown to illustrate that the analytic choices examined here occur in the current literature; this is not an exhaustive systematic search, and each characterization reflects the study's published methods.

| Study | Database | Delirium source | Unable-to-Assess handling | Consequence |
|---|---|---|---|---|
| Zhang et al, 2024 (alcohol use disorder) | MIMIC-IV | CAM-ICU chart item | Excluded ”unable to assess” screens (complete case) | Selection on assess-ability |
| Fu et al, 2024 (body mass index) | MIMIC-IV | CAM-ICU chart item | Excluded ”unable to assess” and missing screens (complete case) | Selection on assess-ability |
| Zhang et al, 2023 (sepsis-associated delirium model) | MIMIC-IV, eICU | CAM-ICU chart item | Excluded ”unable to assess” from training; excluded group later carried a higher predicted delirium burden | Informative exclusion |
| Contreras et al, 2025 (DeLLiriuM) | MIMIC-IV, eICU | Computable RASS-CAM phenotype | No ”unable to assess” label; a positive label requires a positive CAM, so unassessable windows become non-delirium | Silent label construc-tion |

| Study | Database | Delirium source | Unable-to-Assess handling | Consequence |
|---|---|---|---|---|
| Chen et al, 2025 (systematic review, 26 models) | MIMIC-III/IV, eICU, others | Mixed | Missing-data handling ”frequently poorly documented or methodologically unsound” | Review-level |

**eTable 16. Non-English language and Unable to Assess among arousable screens, with ICU-type and admission-era fixed effects.** Cluster-robust logistic regression among arousable screens (RASS -3 or higher; 506,367 screens). The first column reproduces the Figure 3 adjusted model (race and ethnicity, non-English language, arousal, sedation, ventilation, brain injury, age, sex). The second adds ICU-type and admission-era (anchor-year-group) fixed effects to the same model.

| Covariate | Figure 3 model OR (95% CI) | + ICU-type and admission-era fixed effects OR (95% CI) |
|---|---|---|
| Non-English primary language | 3.74 (3.43, 4.08) | 3.94 (3.61, 4.30) |
| Asian | 1.30 (1.12, 1.52) | 1.33 (1.12, 1.57) |
| Hispanic | 1.17 (1.01, 1.36) | 1.12 (0.95, 1.31) |
| Black | 1.13 (1.01, 1.26) | 1.10 (0.99, 1.22) |

Among clearly arousable screens (RASS -2 or higher), the non-English odds ratio with ICU-type and admission-era fixed effects was 4.62 (95% CI, 4.21-5.07), versus 5.01 (95% CI, 4.63-5.43) without them.

The non-English-language association is robust to unmeasured confounding. The E-value for the adjusted odds ratio of 3.74 is 6.95, and the E-value for its lower confidence limit (3.43) is 6.32. An unmeasured confounder would therefore have to be associated with both non-English language and an Unable-to-Assess result by a risk ratio of at least 6.95 each, above and beyond the measured covariates (arousal, sedation, ventilation, brain injury, age, sex, race and ethnicity, ICU type, and admission era), to reduce the observed association to the null; weaker confounding could not. A confounder of this magnitude in the arousable, language-stratified setting is implausible, so unmeasured confounding is unlikely to fully account for the language gap in non-assessment.

**eTable 17. Characteristics of ICU stays removed or made indeterminate by Unable-to-Assess handling.** Complete-case handling drops stays with no scorable (Positive or Negative) screen; missingness-aware handling reports stays with no assessable screen as indeterminate.

| Characteristic | Removed by complete case (all screens UTA; n = 3065) | Retained, ≥1 scorable screen (n = 69,879) | Indeterminate, missingness-aware (n = 2544) |
|---|---|---|---|
| In-hospital mortality, % | 65.2 | 9.8 | 73.5 |
| Mechanical ventilation, ever, % | 61.4 | 28.8 | 67.1 |
| Vasopressor, ever, % | 53.9 | 28.2 | 58.0 |
| Any acute brain injury, % | 45.4 | 18.0 | 49.9 |
| Non-English primary language, % | 15.9 | 9.8 | 10.5 |
| Neuroscience ICU, % | 6.7 | 12.0 | 7.2 |
| ICU length of stay, median, d | 1.3 | 2.1 | 1.3 |

| Characteristic | Removed by complete case (all screens UTA; n = 3065) | Retained, ≥1 scorable screen (n = 69,879) | Indeterminate, missingness-aware (n = 2544) |
| --- | --- | --- | --- |
| Screens at RASS -4 or lower, % | 84.3 | 8.1 | 99.7 |

**eTable 18. Quantitative bias analysis for the UTA-as-negative strategy.** The assumed delirium prevalence among Unable-to-Assess screens (π) was varied from 0% to 100%. For each value, the table reports stay-level delirium prevalence and the baseline-adjusted delirium-mortality odds ratio (mean over 30 random assignments). Of 72,944 stays with complete covariates, 12,982 were not positive under complete-case scoring yet carried at least one UTA screen and were eligible to be reassigned.

| Assumed delirium among UTA screens (π) | Stay-level prevalence, % | Baseline-adjusted mortality OR |
| --- | --- | --- |
| 0% (UTA as negative) | 30.8 | 2.16 |
| 10% | 32.6 | 2.30 |
| 19.3% (≈ observed positive rate) | 34.2 | 2.45 |
| 30% | 36.1 | 2.64 |
| 50% | 39.7 | 3.10 |
| 75% | 44.1 | 3.94 |
| 100% | 48.6 | 5.53 |

The complete-case odds ratio (4.12) is recovered at π of approximately 80%, so the difference between the complete-case and UTA-as-negative estimates corresponds to an unverifiable assumption about delirium status in unassessable patients.

**eTable 19. Inverse-probability-weighted and doubly robust delirium-positive preva-**

**lence among arousable screens.** Target estimand: the positive-screen prevalence among arousable screens (RASS -3 or higher; 506,367 screens) had all such screens been scorable, under missingness at random given observed clinical state. The assessability propensity, P(scored | state), was estimated by cross-fitted logistic regression on RASS, active sedation, neuromuscular blockade, ventilation, hour, neuroscience-ICU indicator, and the eight brain-injury phenotypes; the augmented (doubly robust) estimator adds an outcome model for P(positive | scored, state). Confidence intervals are from a 300-sample patient-cluster bootstrap holding the nuisance models fixed.

| Estimator | Delirium-positive prevalence, % (95% CI) |
|---|---|
| Complete case (scored screens only) | 24.7 |
| UTA recoded as negative | 23.3 |
| Inverse-probability-weighted | 25.6 (25.1, 26.0) |
| Augmented (doubly robust) | 25.6 (25.1, 26.0) |

The maximum stabilized weight was 3.5 (99th percentile, 1.3), so reweighting is well behaved within arousable states. Extending missingness at random into the structurally unassessable screens (RASS -4 or lower) is not defensible, which is why the large between-strategy divergences in the main analysis cannot be removed by adjustment and the state is better preserved than imputed.

**eTable 20. Primary payer and Unable-to-Assess documentation among arousable screens.** Among arousable screens (RASS -3 or higher; 506,367 screens), primary payer was added to the non-English-language model (cluster-robust logistic regression by patient). Absolute Unable-to-Assess (UTA) rates carry Wilson 95% confidence intervals; adjusted odds ratios (aOR) are versus private insurance. P values were Benjamini-Hochberg-corrected over the payer family (3 contrasts) and, separately, over the combined equity family (5 race or ethnicity terms, non-English language, and 3 payer terms). After adding payer, the non-English-language association was essentially unchanged (aOR, 3.65; 95% CI, 3.34-3.99).

| Primary payer | Screens (N) | UTA rate, % (95% CI) | aOR vs private (95% CI) | Significant (BH, equity family) |
|---|---|---|---|---|
| Private | 126,024 | 4.09 (3.98-4.20) | 1 [Reference] | N/A |
| Medicare | 279,520 | 5.97 (5.88-6.06) | 1.08 (0.98-1.19) | No |
| Medicaid | 83,415 | 6.71 (6.54-6.88) | 1.13 (1.01-1.25) | No |
| Other or unknown | 17,408 | 7.49 (7.11-7.89) | 1.40 (1.18-1.66) | Yes |

**eTable 21. External validation of the arousal mechanism in eICU-CRD (208 U.S. hospitals), shown as the mirror image of the MIMIC pattern.** In MIMIC, unassessability is a labeled state, so the fraction of screens marked Unable to Assess rises as arousal falls. In eICU there is no such token, so the same mechanism appears instead as the disappearance of a scored result. Both turn at RASS -4; the small eICU uptick at RASS -5 versus -4 reflects sparse charting in the deepest-sedation stratum. Values are shown for the arousal-depression range (RASS 0 to -5). The eICU column is the percentage of monitored stay-hours containing a scored delirium result (n = 23,968 stays at 56 screening hospitals that also charted RASS); the MIMIC column is the percentage of screens marked Unable to Assess (from the primary analysis).

| RASS | MIMIC: screens marked Unable to Assess (%) | eICU: monitored stay-hours with a scored result (%) |
|---|---|---|
| 0 | 2.0 | 29.1 |
| -1 | 7.7 | 18.7 |
| -2 | 13.2 | 16.3 |
| -3 | 19.9 | 19.0 |
| -4 | 97.8 | 9.2 |
| -5 | 99.5 | 10.3 |

In eICU, a scored delirium result was present in 24.2% of arousable stay-hours (RASS -3 or higher) versus 9.7% of deeply sedated ones (RASS -4 or -5), and only 2.1% of scored screens occurred at RASS -4 or below; the odds of a scored result rose 1.24-fold per one-point increase in RASS (95% CI, 1.24 to 1.25). The arousal mechanism therefore reproduces in an independent multicenter cohort despite the opposite documentation convention.

### S3. Supplementary Figures

**eFigure 1. Unscreenability probe.** Receiver operating characteristic curves for predicting whether a delirium screen is recorded as Unable to Assess, from the patient's structural state only (logistic regression and gradient boosting; out-of-fold predictions, patient-grouped 5-fold cross-validation). High discrimination indicates that unassessability is a near-deterministic function of arousal and sedation, that is, informative missingness.

*(File: eFigure1_probe_roc.pdf / .png)*

### S4. Code and Data Availability

MIMIC-IV version 3.1 and the eICU Collaborative Research Database version 2.0 are available to credentialed users through PhysioNet (https://physionet.org/content/mimiciv/3.1/ and https://physionet.org/content/eicu-crd/2.0/). Analysis code that reproduces every number, table, and figure in this report will be made publicly available at https://github.com/Alon-Gorenshtein/study_delirium_unscreenable (archived on Zenodo on publication). No patient-identifiable data are contained in this Supplementary Information.
